# Branched chain amino acids and risk of breast cancer

**DOI:** 10.1101/2020.08.31.20185470

**Authors:** Oana A. Zeleznik, Raji Balasubramanian, Yumeng Ren, Deirdre K. Tobias, Bernard Rosner, Cheng Peng, Alaina M. Bever, Lisa Frueh, Clary B. Clish, Samia Mora, Frank Hu, A. Heather Eliassen

**Affiliations:** Channing Division of Network Medicine, Brigham and Women’s Hospital and Harvard Medical School, Boston, MA, USA; Department of Biostatistics & Epidemiology, University of Massachusetts - Amherst, Amherst, MA, USA; Department of Epidemiology, Harvard T.H. Chan School of Public Health, Boston, MA; Division of Preventive Medicine, Department of Medicine, Brigham and Women’s Hospital and Harvard Medical School, Boston, MA; Broad Institute of Massachusetts Institute of Technology and Harvard, Cambridge, MA, USA; Department of Nutrition, Harvard T.H. Chan School of Public Health, Boston, MA, USA

## Abstract

Circulating branched chain amino acid (BCAA) levels reflect metabolic health as well as dietary intake and have been linked to some cancers. Associations with breast cancer are unclear.

We evaluated the association between circulating BCAA levels and risk of breast cancer in a prospective nested case-control study (1,997 cases, 1,997 controls) within the Nurses’ Health Study (NHS) and NHSII. Two-thirds of women in NHS (592 cases) donated two blood samples collected 10 years apart. We used conditional logistic regression to estimate odds ratios (OR) and 95% confidence intervals (CI) of breast cancer risk in multivariable models which included BMI at age 18 and adulthood weight gain, in addition to other risk factors. We conducted an external validation with secondary analyses in the Women’s Health Study (WHS) (1,297 cases).

Among NHSII participants (predominantly premenopausal at blood collection), elevated circulating BCAA levels were associated with suggestively lower breast cancer risk (e.g., isoleucine highest vs. lowest quartile, multivariable OR (95% CI)= 0.86 (0.65-1.13), p-trend=0.20), with significant linear trends among fasting samples (e.g., isoleucine OR (95% CI)=0.74 (0.53-1.05), p-trend=0.05). In contrast, among postmenopausal women, proximate measures (within 10y from blood draw) were associated with increased breast cancer risk (e.g., isoleucine highest vs. lowest quartile multivariable OR (95% CI)=1.63 (1.12-2.39), p-trend=0.01), with slightly stronger associations among fasting samples (OR (95% CI)=1.73 (1.15-2.61), p-trend=0.01). Distant measures (10-20y since blood draw) were not statistically significantly associated with risk (OR (95% CI)=1.15 (0.87-1.52), p-trend=0.35). We did not observe significant heterogeneity by ER status or BMI. In the WHS, a suggestive positive association was observed for distant measures of leucine among postmenopausal women: OR (95% CI)=1.31 (0.97-1.75), p-trend=0.05.

Although elevated circulating BCAA levels were associated with lower breast cancer risk among premenopausal NHSII women and higher risk of postmenopausal breast cancer in NHS when assessed within 10 years of diagnosis, independent of established risk factors, including adiposity, results were not validated in WHS. Additional independent studies are needed to reassess and understand the complex relationship between BCAAs, menopausal status and timing, and risk of breast cancer.

**Statement of significance:** Elevated circulating BCAA levels were associated with lower breast cancer risk among premenopausal NHSII women and higher risk of postmenopausal breast cancer in NHS when assessed within 10 years of diagnosis, independent of established risk factors, including adiposity.

## Introduction

Breast cancer is the most common malignancy in women, with more than 250,000 women diagnosed annually in the United States^1^. Epidemiologic studies have identified modifiable risk factors associated with breast cancer, including increased BMI and low physical activity in postmenopausal women^2^. However, BMI is inversely associated with premenopausal breast cancer. These findings indicate that poor metabolic health may be associated with the development or progression of breast cancer, although mechanisms and explanations for the variation by menopausal status remain unclear.

The branched chain amino acids (BCAA) leucine, valine, and isoleucine are essential amino acids obtained from diet, and important metabolites involved in cell-signaling pathways and muscle protein synthesis^3^. Elevated plasma BCAA concentrations are strongly positively correlated with BMI and insulin resistance, and appears to be a marker of dysfunctional metabolism^4^. In light of recent human studies linking plasma BCAA concentrations with risk of cancer and diabetes^5,6^, plasma BCAA concentration is a potential biomarker of interest in understanding breast cancer mechanisms in relation to metabolism and diet.

Hypotheses regarding the association between plasma BCAAs and cancer development are multifaceted: increased BCAA in plasma may play a causal role by directly activating pathways related to cancer growth and metastasis, or elevated BCAA may be the consequence of dysregulated metabolism or cancer-related muscle wasting^6^. Current evidence suggesting a link between BCAAs and breast cancer includes in vitro studies showing up-regulation of breast cancer cell growth in response to BCAA metabolism pathways^7,8^, and increased plasma BCAA concentration in human breast cancer patients at diagnosis or post-diagnosis^8,9^. Whether elevated BCAA concentration is associated with breast cancer incidence, and how BCAA is differentially related to breast cancer by menopausal status, remains unknown.

To date, two epidemiologic studies from the prospective EPIC cohort reported no associations between circulating plasma BCAA levels and breast cancer risk among a mix of pre- and postmenopausal women^10,11^, while another smaller study found a positive association between plasma valine and breast cancer risk^12^. Only one study conducted analyses by menopausal status and found positive associations between allo-isoleucine and postmenopausal breast cancer^13^; however this study included only metabolites correlated with BMI in the study population, resulting in the exclusion of isoleucine and leucine from the analysis. We conducted a nested case-control study within the Nurses’ Health Study (NHS) and NHSII to investigate the association between plasma BCAA levels with risk of breast cancer. In secondary analyses, we conducted a validation analysis in the Women’s Health Study (WHS)

## Methods

### Study Population

In 1976, 121,701 female registered nurses aged 30-55y returned a mailed questionnaire and were enrolled in the NHS^14^. Participants have been followed biennially since 1976 with questionnaires collecting information on reproductive history, lifestyle factors, diet, medication use, and new disease diagnoses. The NHSII was begun in 1989 with 116,429 female registered nurses aged 25-42y. Participants have been followed biennially using similar questionnaires as NHS.

In 1989-1990, 32,826 NHS participants aged 43-69y contributed blood samples, as previously described^15^. Briefly, each woman arranged to have her blood drawn and shipped it overnight to our laboratory where we processed it and archived aliquots of white blood cell, red blood cell, and plasma in liquid nitrogen freezers (≤-130°C). In 2000-2002, 18,473 of these women aged 53-80y donated a second blood sample using a similar protocol. In the NHSII, 29,611 women aged 32-54y donated blood and urine samples in 1996-1999. Of these, 18,521 women who had not used oral contraceptives, been pregnant or breastfed in the previous six months provided samples timed within the menstrual cycle, targeting the early follicular (days 3 to 5 of the cycle) and mid-luteal (7 to 9 days prior to expected start of next cycle) phases. The remaining women donated a single untimed sample. Follicular plasma was separated and frozen by the participants and returned with the luteal sample; luteal and untimed samples were collected and shipped following the same protocol as NHS. Follow-up in the blood subcohorts is high (NHS 97% in 2010; NHSII 96% in 2011).

The study protocol was approved by the institutional review boards of the Brigham and Women’s Hospital and Harvard T.H. Chan School of Public Health, and those of participating registries as required. The return of the self-administered questionnaire and blood sample was considered to imply consent.

Details about the WHS can be found in the supplementary materials. Associations among WHS women were evaluated restricting to fasting participants, by menopausal status at blood collection and among postmenopausal women by time between blood collection and diagnosis.

### Case and Control Selection

Incident cases of breast cancer were identified after blood collection among women who had no reported cancer (other than non-melanoma skin). Cases were diagnosed between 2000 and 2010 (NHS) or 1999-2011 (NHSII). Overall, 940 cases were included in the NHS, 592 of whom had donated blood samples in both the first (1989-1990) and second (2000-2002) blood collections. In the NHSII (1996-1999), 1,057 cases were included. Breast cancer cases were reported (invasive cases: NHS=748; NHSII=780) and confirmed by medical record reviews (NHS=914; NHSII=1015) or verbally by the nurse (NHS=26; NHSII=42). Given the high confirmation rate by medical record for breast cancer in this cohort (99%), all cases are included in this analysis.

In NHS, one control was matched per case by the following factors (at both collections for subjects with 2 samples): age (+/- 2y), menopausal status and postmenopausal hormone therapy (HT) use at blood collection and diagnosis (premenopausal, postmenopausal and not taking HT, postmenopausal and taking HT, and unknown), and month (+/- 1mo), time of day (+/- 2h), and fasting status at blood collection (<8 h after a meal or unknown; >8h). In NHSII, one control was matched per case on the same factors as NHS, with the addition of race/ethnicity (African American, Asian, Hispanic, Caucasian, other) and luteal day (+/- 1d; timed samples only).

### Laboratory Assays

#### BCAAs measurement

In the NHS/NHSII, BCAAs were assayed through a metabolomic profiling platform at the Broad Institute using a liquid chromatography tandem mass spectrometry (LC-MS) method designed to measure polar metabolites such as amino acids, amino acids derivatives, dipeptides, and other cationic metabolites^16-18^. Pooled plasma reference samples were included every 20 samples and results were standardized using the ratio of the value of the sample to the value of the nearest pooled reference multiplied by the median of all reference values for the metabolite. Hydrophilic interaction liquid chromatography (HILIC) analyses of water-soluble metabolites in the positive ionization mode were conducted using an LC-MS system comprised of a Shimadzu Nexera X2 U-HPLC (Shimadzu Corp.; Marlborough, MA) coupled to a Q Exactive mass spectrometer (Thermo Fisher Scientific; Waltham, MA). Metabolites were extracted from plasma (10 μL) using 90 μL of acetonitrile/methanol/formic acid (74.9:24.9:0.2 v/v/v) containing stable isotope-labeled internal standards (valine-d8, Sigma-Aldrich; St. Louis, MO; and phenylalanine-d8, Cambridge Isotope Laboratories; Andover, MA). The samples were centrifuged (10 min, 9,000 x g, 4°C), and the supernatants were injected directly onto a 150 x 2 mm, 3 μm Atlantis HILIC column (Waters; Milford, MA). The column was eluted isocratically at a flow rate of 250 μL/min with 5% mobile phase A (10 mM ammonium formate and 0.1% formic acid in water) for 0.5 minute followed by a linear gradient to 40% mobile phase B (acetonitrile with 0.1% formic acid) over 10 minutes. MS analyses were carried out using electrospray ionization in the positive ion mode using full scan analysis over 70-800 m/z at 70,000 resolution and 3 Hz data acquisition rate. Other MS settings were: sheath gas 40, sweep gas 2, spray voltage 3.5 kV, capillary temperature 350°C, S-lens RF 40, heater temperature 300°C, microscans 1, automatic gain control target 1e6, and maximum ion time 250 ms. Metabolite identities were confirmed using authentic reference standards or reference samples. NHS samples were run together, followed by NHSII samples. The coefficients of variation (CV) among blinded quality control samples (N=638) were <15% (isoleucine=7.3-11.4%, leucine=7.0-12.1%, valine=6.2-9.8%). BCAAs were not affected by delayed processing of blood samples and showed good within person stability over 1-2 years (intra-class correlation≥0.55)^19^.

In the WHS, baseline blood samples from a subset of 28,345 women were shipped on ice via overnight courier to the central laboratory where they were processed and stored at - 170 degrees C in vapor liquid nitrogen. Aliquots of the EDTA plasma samples were shipped on dry ice blinded to outcome status to LipoScience, Inc, now LabCorp® (Raleigh, NC). Isoleucine, leucine, and valine were measured by proton nuclear magnetic resonance (^1^H NMR) spectroscopy using a 400MHz NMR platform, as described for the NMR LipoProfile® IV test.^20,21^ The derived BCAA signal amplitudes were converted to μmol/L, with intra- and inter-assay coefficients of variation of isoleucine 5.9-6.1%, leucine 4.5-4.9%, and valine 1.5-2.1 %.

#### Gene expression

Archived formalin-fixed paraffin-embedded breast cancer tissue blocks were collected and assembled into tissue microarrays (TMAs)^22^. RNA was extracted from multiple cores of 1 or 1.5 mm taken from formalin fixed paraffin-embedded (FFPE) tumor (n=1-3 cores) and normal-adjacent (n=3-5 cores) tissues using the Qiagen AllPrep RNA isolation kit^23^. Normal-adjacent tissues were obtained >1 cm away from the edge of the tumor. A detailed protocol has been published previously^24-26^ (microarray accession number: GSE115577). In brief, we profiled transcriptome-wide gene expression using Affymetrix Glue Grant Human Transcriptome Array 3.0 (hGlue 3.0) and Human Transcriptome Array 2.0 (HTA 2.0) microarray chips (Affymetrix, Santa Clara, CA, USA). We used robust multi-array average to perform normalization (RMA; Affymetrix Power Tools (ATP)), log-2 transformed the data, and conducted sample quality control with Affymetrix Power Tools probeset summarization based metrics^24,25^. A total of 882 tumor tissues from invasive breast cancer cases passed quality control. Among these, 120 NHSII (68% premenopausal at blood collection) samples and 101 NHS samples (all postmenopausal) had gene expression and BCAA levels measured. For genes that were mapped by multiple probes, we selected the most variable probe to represent the gene. Our current analyses included 17,791 (70%) genes that were profiled in both platforms. Technical variabilities by batch were controlled using *ComBat*, an empirical Bayes method used to control for known batch effects^27^. Genes with low expression (<25^th^ percentile) were removed from the analyses.

#### Estradiol and C-peptide

Estradiol assays were performed at Quest Diagnostics (San Juan Capistrano, CA) as previously described^28^ by radioimmunoassay following extraction and celite column chromatography (NHSII, luteal measures N=278 cases, 280 controls; follicular measures N=268 cases, 264 controls) or at the Mayo Clinic (Rochester, MN) by LC-MS/MS as previously described^29^ (NHS distant measures, N=145 cases, 143 controls; NHS proximate measures, N=119 cases, 115 controls). C-peptide was assessed using ELISA (Diagnostic Systems Laboratory, Webster, TX) in the laboratory of Dr. Michael Pollak (McGill University, Montreal, Quebec, Canada) (NHSII, N=290 cases, 289 controls; NHS distant collection measures N=202 cases, 205 controls; NHS proximate collection measures N=124 cases, 120 controls).

#### Covariate Information

Data on breast cancer risk factors, including anthropometric measures, reproductive history, and lifestyle factors, were collected from questionnaires administered biennially and at the time of blood collections. Case characteristics, including invasive vs. *in situ*, histologic grade, estrogen and progesterone receptor (ER, PR), and human epidermal growth factor receptor 2 (HER2) status, were extracted from pathology reports. As previously described^30^, immunohistochemical results for ER, PR, and HER2, read manually by a study pathologist, were included for cases with available tumor tissue included in TMAs.

#### Statistical Analysis

BCAA values were log transformed and standardized to mean = 0 and standard deviation (SD) =1 within each cohort and blood collection separately. In order to estimate the association between BCAAs as a group and risk of breast cancer we calculated the sum of all three BCAAs (total BCAAs) and considered it an additional exposure in our analyses.

We estimated within-person stability over 10 years by calculating intra-class correlation (ICC) using mixed liner models and Spearman correlation coefficients among participants who donated two blood samples approximately 10 years apart.

We used linear regression models to investigate factors associated with circulating BCAA levels, including fasting status, date of blood collection, individual dietary BCAA intake and caloric intake from validated food frequency questionnaires from around the time of blood collection, cohort, age, race, BMI, physical activity, alcohol consumption, smoking status, alternative healthy eating index, case-control status among NHS/NHSII (N=9,112) women from nested case-control studies conducted across 14 endpoints (e.g., breast cancer, colon cancer, stroke, glaucoma).

Conditional logistic regression was used to evaluate the associations between BCAAs and risk of breast cancer in each cohort separately. We estimated odds ratios (OR) and 95% confidence intervals (95%CI) across quartiles (based on the distribution in controls) of BCAA levels and used quartile medians (calculated based on the control distribution) to estimate linear trend p-values. In a sensitivity analysis, we compared conditional to unconditional logistic regression adjusted for matching factors and obtained similar results (data not shown). Based on these findings, we used unconditional logistic regression in analyses stratified by BMI and ER status.

In multivariable models, we adjusted for established breast cancer risk factors: BMI at age 18, weight change from age 18 to time of blood draw, age at menarche, parity and age at first birth, family history of breast cancer, history of benign breast disease, physical activity, alcohol consumption, exogenous hormone use, and breastfeeding history. In a separate analysis among NHS participants, we cross-classified participants based on the median BCAA levels among controls at the two blood collections and categorized women at first and second blood collection as low/low, low/high, high/low, or high/high. In the WHS, we used Cox proportional hazards regression models with follow-up from the date of randomization to date of first invasive cancer diagnosis, death, or December 31, 2018. We assessed heterogeneity between NHS/NHSII and WHS using the DerSimonian-Laird estimator ^31^, and based on these findings, meta-analyzed individual cohort results using a fixed or random effects approach.

Using a competitive gene set testing procedure, Correlation Adjusted Mean Rank (CAMERA), on the gene expression data, we explored functional enrichment of biological pathways associated with BCAA^32^. We chose the 50 "hallmark” gene sets from the Molecular Signature Database (MSigDB)^33^. We controlled for age and year of diagnosis, BMI at blood draw, alcohol consumption, menopausal status, menopausal hormone use, ER status, race, as well as matching factors. We chose an inter-gene correlation of 0.01.

We conducted sensitivity analyses, including restricting to fasting samples (>8h since last meal), restricting to premenopausal or postmenopausal women at blood collection, adjusting for BMI at the time of the blood collection instead of BMI at age 18 and weight change between age 18 and blood collection, and adjusting for C-peptide (a marker of insulin production) and estradiol in individual models.

Analyses were conducted using R version 3.6.0, R version 3.1.4 and SAS Version 9.3 software (SAS Institute, Cary, NC)

## Results

In total, 1997 matched case control pairs were analyzed in this study (Table 1, Figure 1). There were 1057 cases and 1057 matched controls from NHSII, with a mean age at blood collection of 45 years (Table 1). These women were predominantly premenopausal (80%) at the time of blood collection (Figure 1). The remaining cases and controls were participants of the NHS cohort: 940 cases and their matched controls donated a blood sample during the first blood collection (1989-1990) and of these, 592 cases and their matched controls donated a second blood sample (2000-2002). NHS participants were on average 55 years old at the time of first blood collection (distant measure) and predominantly postmenopausal (62%). At the second collection (proximate measure), participants were on average 66 years old and postmenopausal (98%). The mean times between blood collection and diagnosis were: 8 years for NHSII, 15 years for NHS distant measure, and 4 years for NHS proximate measure.

**Figure 1.**
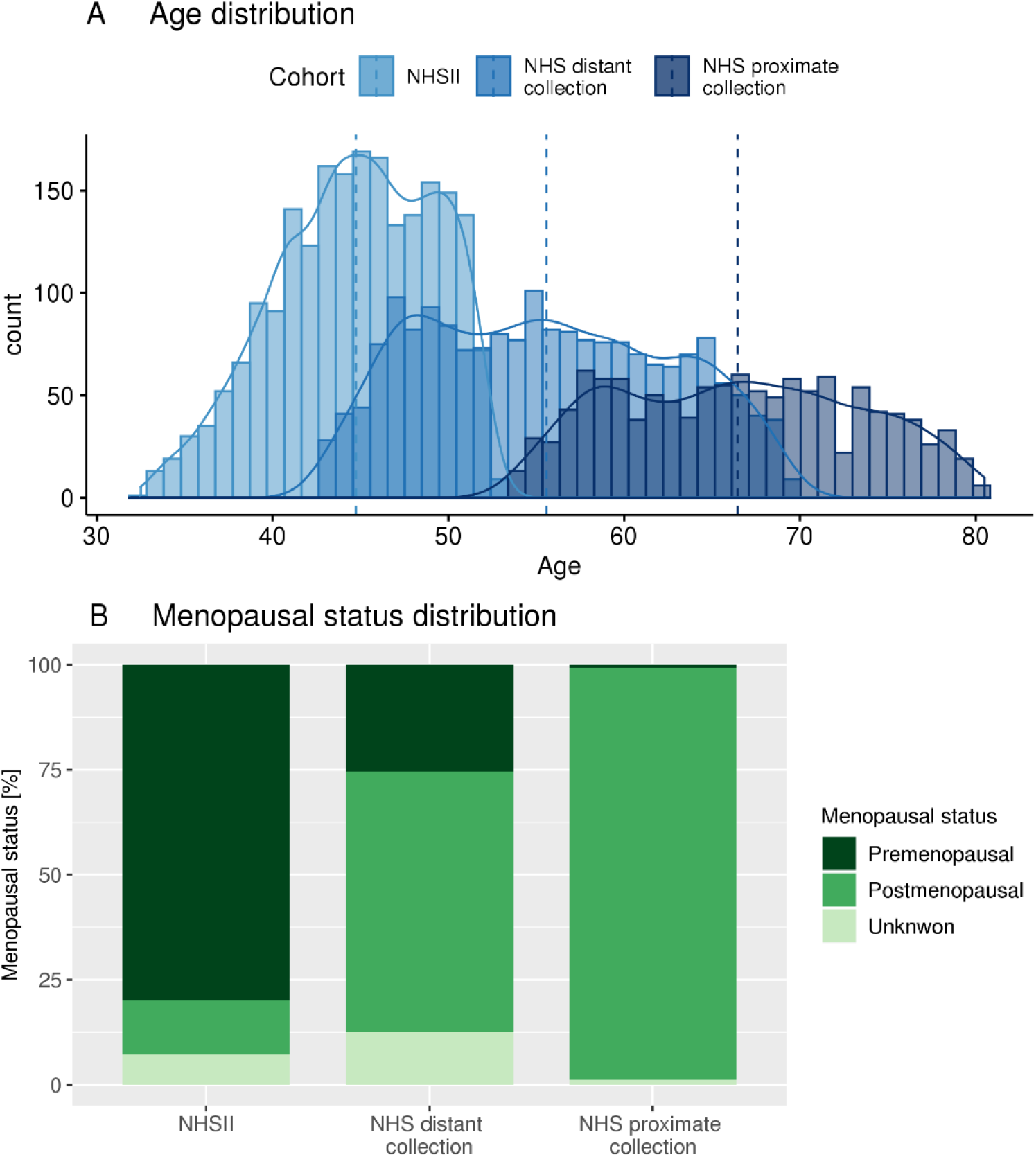
Age and menopausal status distribution at blood collection.

**Table 1:** Characteristics of breast cancer cases and matched controls in the Nurses’ Health Studies.

|  | NHSII |  | NHS distant collection<sup>1</sup> |  | NHS proximate collection<sup>2</sup> |  |
| --- | --- | --- | --- | --- | --- | --- |
|  | Cases | Controls | Cases | Controls | Cases | Controls |
|  | N=1057 | N=1057 | N=940 | N=940 | N=592 | N=592 |
| Age at blood collection<sup>^</sup>, y | 44.7 (4.5) | 44.8 (4.4) | 55.5 (6.9) | 55.6 (6.9) | 66.4 (6.9) | 66.5 (6.8) |
| Time between blood collection and diagnosis, y | 8.0 (4.4) |  | 14.6 (3.0) |  | 4.0 (2.6) |  |
| Age at menarche, y | 12.4 (1.3) | 12.5 (1.4) | 12.5 (1.4) | 12.6 (1.4) | 12.5 (1.4) | 12.6 (1.4) |
| Parity and age at first birth, %: |  |  |  |  |  |  |
| Nulliparous | 21.1 | 18.4 | 9.6 | 8.0 | 8.6 | 5.9 |
| 1-2 children <25y | 14.7 | 15.9 | 13.5 | 14.1 | 13.0 | 15.9 |
| 1-2 children ≥25y | 39.2 | 34.9 | 20.1 | 20.6 | 20.4 | 19.3 |
| 3+ children <25y | 11.3 | 16.6 | 35.5 | 35.5 | 36.5 | 38.3 |
| 3+ children ≥25y | 13.8 | 14.2 | 21.3 | 21.7 | 21.5 | 20.6 |
| Family history of breast cancer, % | 17.4 | 10.8 | 14.6 | 10.7 | 22.5 | 14.2 |
| Personal history of benign breast disease, % | 22.1 | 15.6 | 45.9 | 37.8 | 62.5 | 54.7 |
| BMI at age 18, kg/m<sup>2</sup> | 20.8 (2.9) | 21.1 (3.1) | 21.1 (2.7) | 21.3 (3.0) | 21.0 (2.6) | 21.3 (3.0) |
| Weight change between age 18 and blood collection, kg | 11.6 (12.0) | 12.6 (13.2) | 12.3 (10.9) | 10.6 (11.2) | 15.5 (12.7) | 13.8 (12.7) |
| Physical activity, MET-hrs/wk | 18.0 (15.3) | 18.1 (15.5) | 15.4 (18.8) | 15.9 (17.6) | 17.7 (14.8) | 19.0 (17.7) |
| Alcohol consumption, g/day | 3.8 (6.9) | 3.3 (5.6) | 6.9 (9.9) | 5.9 (8.2) | 6.7 (9.2) | 5.8 (7.7) |
| Past/current exogenous hormone use*, % | 86.3 | 86.7 | 68.1 | 68.2 | 80.6 | 81.2 |
| Ever breastfed, % | 63.1 | 65.0 | 64.3 | 62.0 | 67.4 | 64.4 |
| Menopausal status at blood collection<sup>^</sup>, % |  |  |  |  |  |  |
| Premenopausal | 80.2 | 79.7 | 25.4 | 25.5 | 0.5 | 0.8 |
| Postmenopausal | 12.7 | 13.1 | 61.9 | 61.9 | 98.1 | 98.1 |
| Unknown | 7.1 | 7.3 | 12.7 | 12.6 | 1.4 | 1.0 |
| Menopausal status at diagnosis<sup>^</sup>, % |  |  |  |  |  |  |
| Premenopausal | 42.0 | 42.2 | 1.3 | 1.3 | 1.4 | 1.0 |
| Postmenopausal | 46.4 | 47.1 | 97.3 | 98.1 | 97.8 | 98.3 |
| Unknown | 11.6 | 10.7 | 1.4 | 0.6 | 0.8 | 0.7 |
| Fasting (>8h) at blood collection<sup>^</sup>, % | 68.7 | 74.7 | 66.6 | 72.7 | 87.0 | 92.4 |
| Caucasian<sup>^</sup>, % | 97.2 | 98.4 | 98.3 | 98.8 | 98.6 | 99.5 |

<sup>1</sup> NHS first blood collection; <sup>2</sup> NHS second blood collection <sup>^</sup> matching factor; \* oral contraceptive or menopausal hormone therapy

WHS included 54% postmenopausal and 46% premenopausal women at the time of sample collection. The mean time between sample collection and diagnosis was similar to NHS and NHSII: 6 years for postmenopausal cases with proximate samples collection, 16 years for postmenopausal cases with distant blood collection and 5 years for premenopausal women at blood collection. Demographics of WHS women were similar to NHS women; exceptions include lower family history of breast cancer (Table 1a).

**Table 1a:**
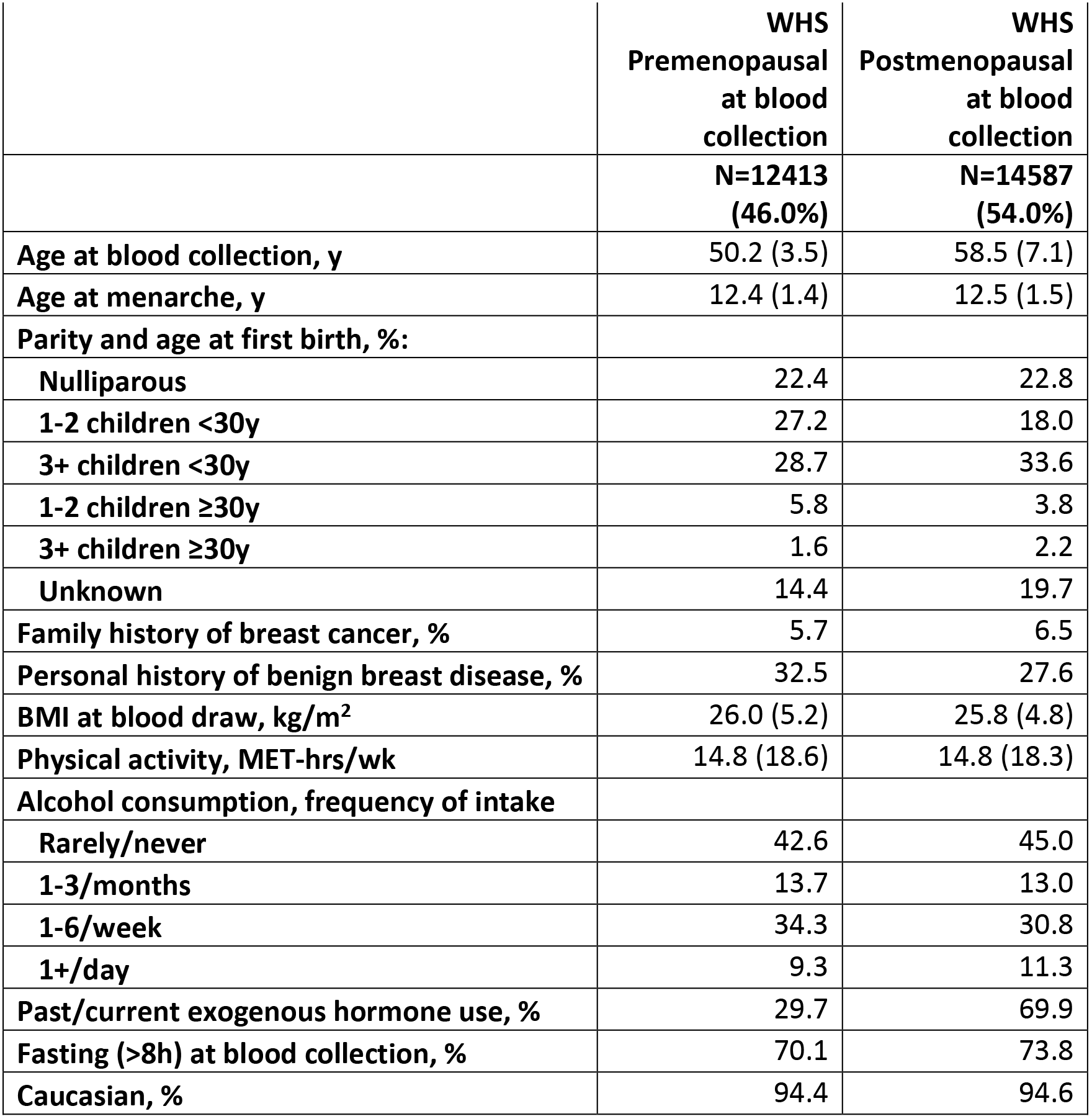
Characteristics of the Women’s Health Study.

BCAA levels were reasonably stable over 10 years among the subset of women with repeated measures (N=592; ICC isoleucine=0.45, leucine=0.44, valine=0.48). Several factors were significantly associated with circulating BCAA levels (Table 2). Factors significantly positively associated with BCAA levels included: dietary intake of BCAAs, BMI, non-fasting blood collection, and Asian Americans compared to Caucasians. Factors significantly inversely associated with BCAA levels included: alcohol consumption and diet quality (alternative healthy eating index^34^).

**Table 2:**
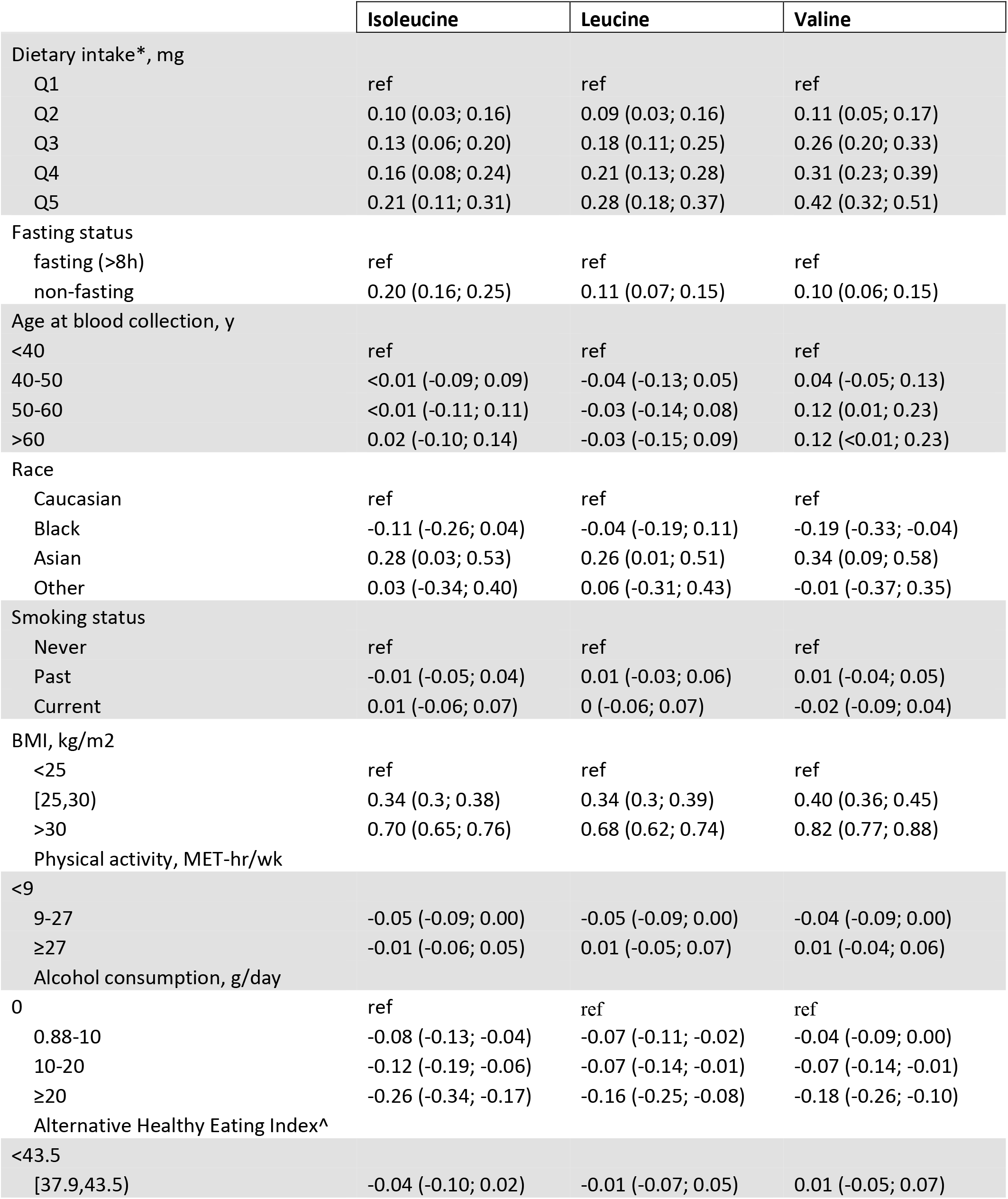
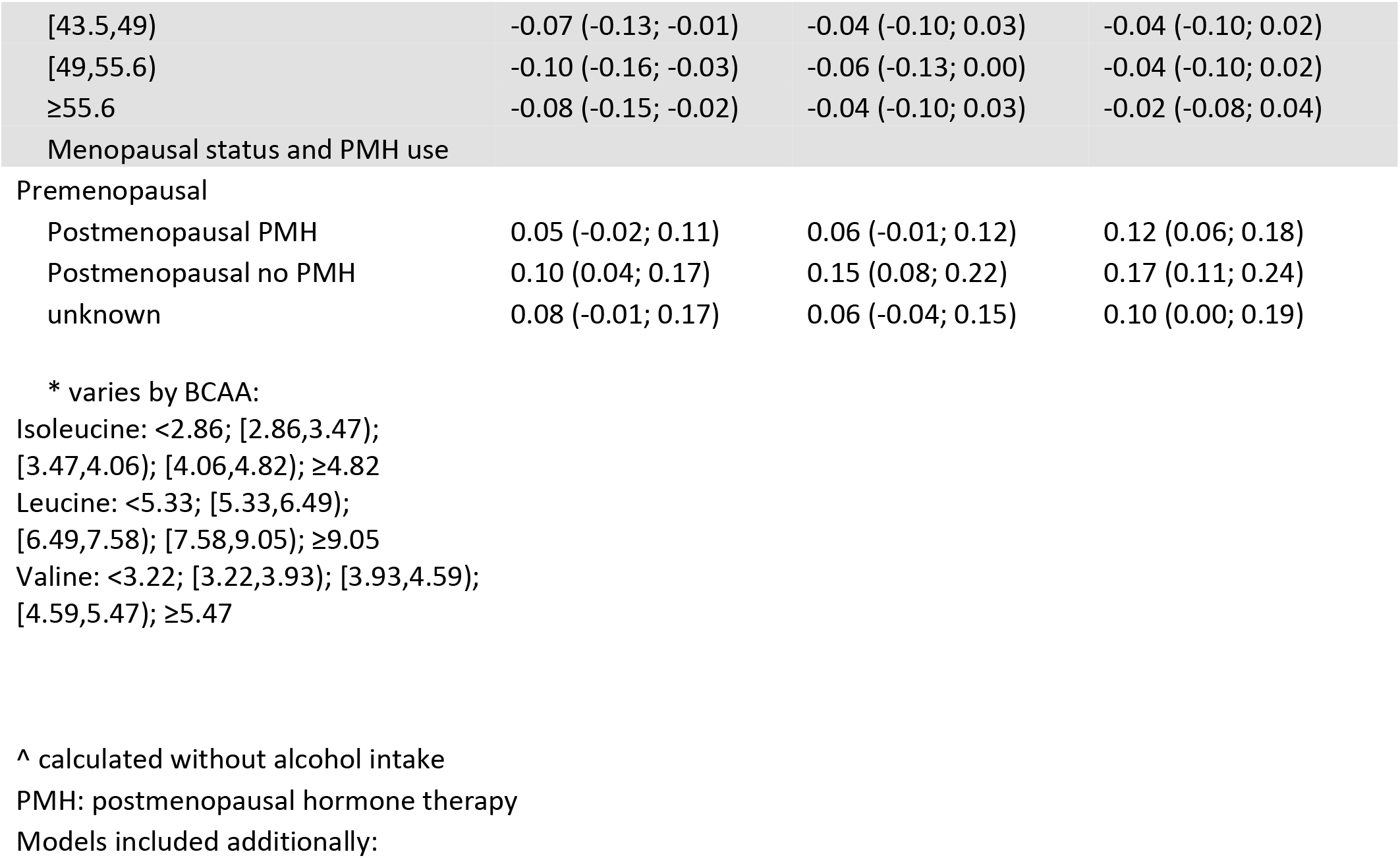
Effect estimates β and 95% confidence intervals (95%CI) for factors potentially associated with 1 SD increase in probit transformed circulating BCAA levels among 9,112 NHS/NHSII women.

Among premenopausal women at blood collection, individual BCAAs were inversely associated with risk of breast cancer (simple model) (e.g., isoleucine highest vs. lowest quartile OR (95% CI)=0.76 (0.59-0.99, p-trend=0.02); Table 3), with significant linear trends. These associations were attenuated and no longer statistically significant when we adjusted for breast cancer risk factors (e.g., isoleucine highest vs. lowest quartile OR (95% CI)=0.86 (0.65-1.13), p-trend=0.20). Associations were similar for leucine (OR (95%CI)=0.77 (0.58-1.01)) and valine (0.82(0.62-1.08)). We observed stronger associations among those with fasting samples only (top vs. bottom quartile OR (95% CI): isoleucine=0.74 (0.53-1.05), p-trend=0.05; leucine=0.66 (0.47-0.94), p-trend=0.04; valine=0.74 (0.53-1.04), p-trend=0.08). Associations with the total BCAAs followed a similar pattern, but were attenuated compared to the individual BCAAs: OR (95%)=0.79 (0.56-1.11), p-trend=0.12. We observed slightly stronger associations for leucine and valine when we further restricted to premenopausal women at blood collection (OR (95%CI); leucine=0.61 (0.40-0.92), p-trend=0.04; valine=0.66 (0.45-0.98), p-trend=0.02; data not shown). Associations were similar and slightly stronger for valine when we restricted to women who were premenopausal at the time of diagnosis (valine=0.60 (0.32-1.11), p-trend=0.14; data not shown).

**Table 3:**
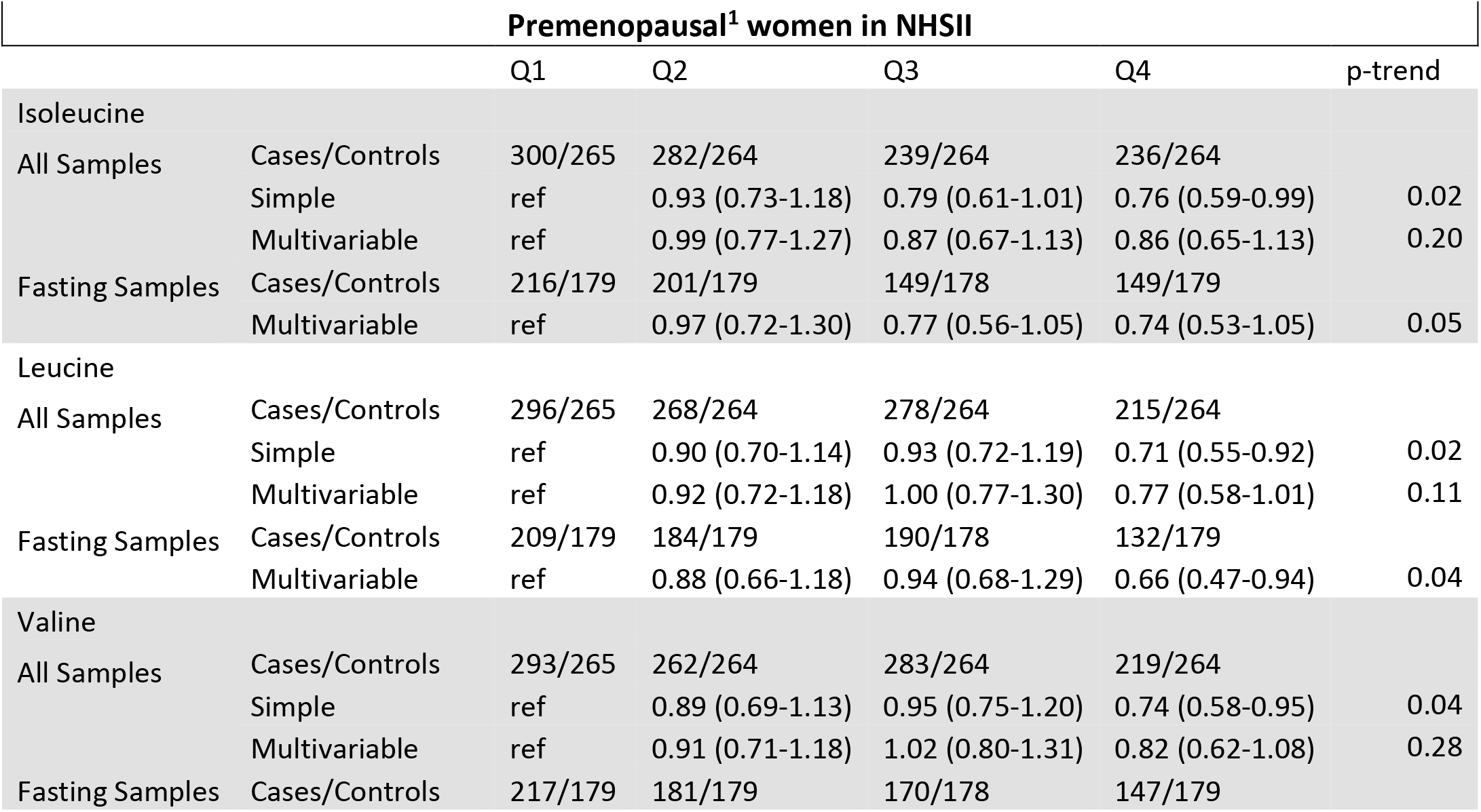
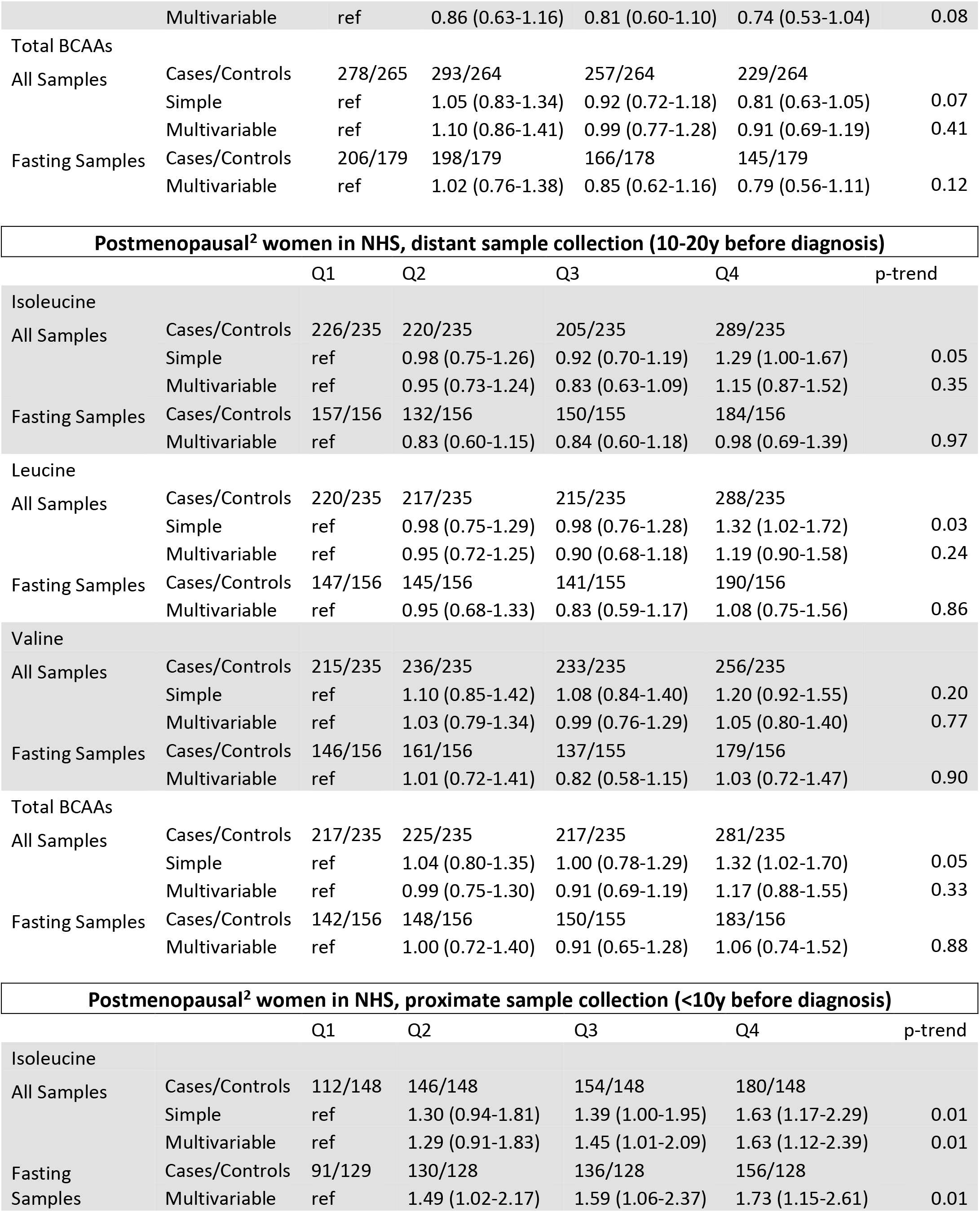
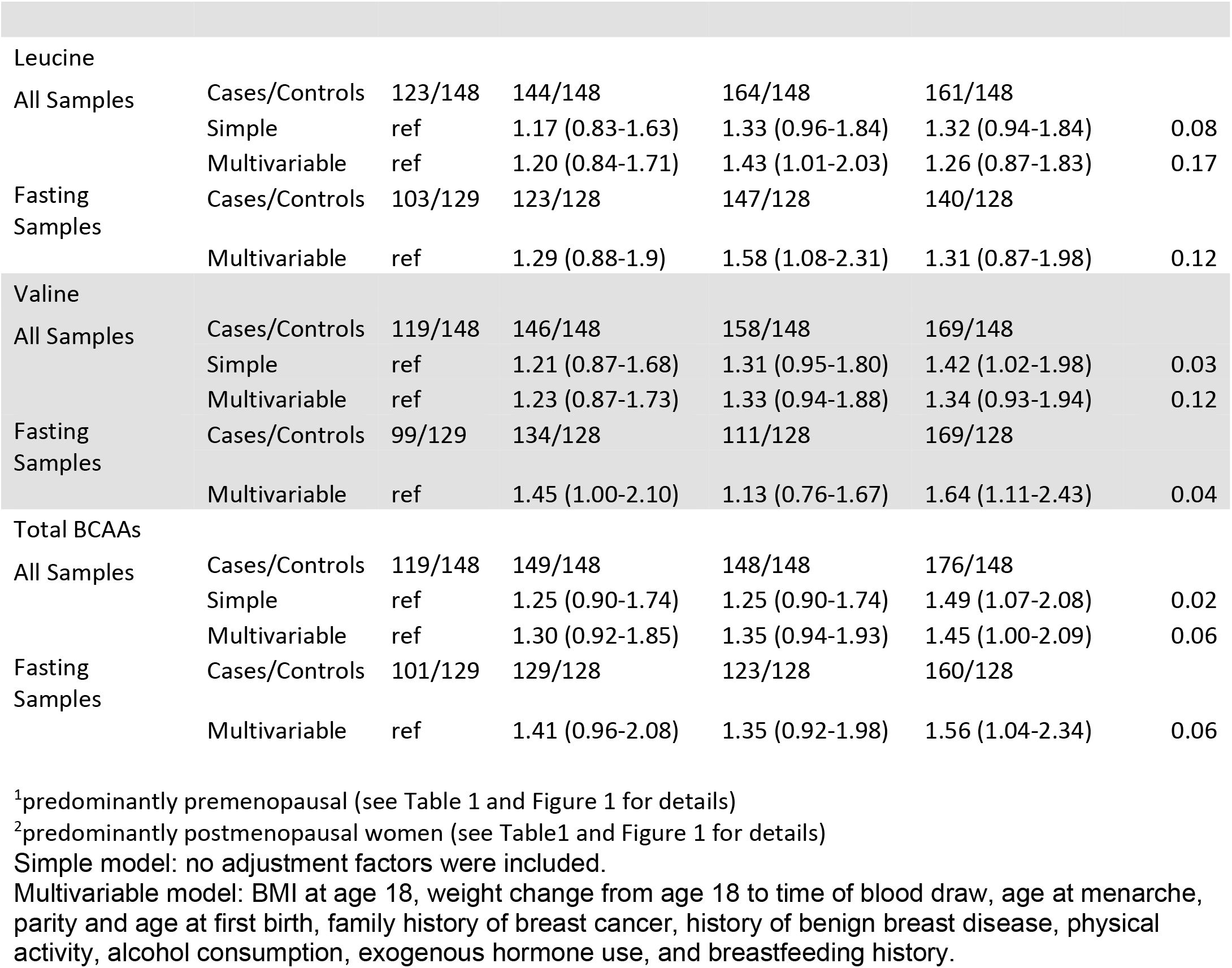
Odds ratios (OR) and 95% confidence intervals (CI) of breast cancer according to quartiles of plasma branched chain amino acids in premenopausal and postmenopausal women.

Among postmenopausal women, we observed positive associations between distant (10-20y before diagnosis) measures of isoleucine and leucine and breast cancer risk in the simple model; however, these were attenuated and no longer significant with multivariable adjustment (e.g., isoleucine OR (95% CI)=1.15 (0.87-1.52), p-trend=0.35). Individual BCAAs from proximate samples were positively associated with breast cancer risk and similar between the simple and multivariable models (e.g., isoleucine multivariable OR (95% CI)=1.63 (1.12-2.39), p-trend=0.01). Weaker associations were observed for leucine (OR (95% CI)=1.26 (0.87-1.83), p-trend=0.17) and valine (1.34 (0.93-1.94), p-trend=0.12). Associations were stronger, with significant linear trends (except for leucine), when restricted to fasting samples (isoleucine=1.73 (1.15-2.61), p-trend=0.01; leucine=1.31 (0.87-1.98), p-trend=0.12; valine=1.64 (1.11-2.43), p-trend=0.04). Association with total BCAAs followed the same pattern as the individual BCAAs: OR (95%)=1.56 (1.04-2.34), p-trend=0.06 in the multivariable model among fasting women. A significant interaction with menopausal status (p<0.03) was observed when we pooled NHSII and NHS women with proximate measures.

Individual and total BCAAs were not associated with risk of breast cancer among either all or fasting WHS premenopausal or postmenopausal women with distant or proximate blood collections in the simple or the multivariable models. For example, among fasting postmenopausal women with proximate measures multivariable OR(95%CI) for isoleucine = 0.97(0.72-1.31), p-trend=0.71 (Table 3a). A suggestive positive association was observed for leucine and risk of breast cancer among all and when restricting to fasting postmenopausal women with distant sample collection: multivariable OR(95%CI)=1.31 (0.97-1.75), p-trend=0.05. There were too few premenopausal women at diagnosis to examine these associations in WHS (N=36). We observed significant heterogeneity between the cohorts among postmenopausal women with proximate blood collection (except for leucine). Among premenopausal women at blood collection, only leucine shown significant heterogeneity between NHSII and WHS women. We observed no significant associations between individual and total BCAA levels and risk of breast cancer when meta-analyzing NHS/NHSII and WHS results (e.g., isoleucine top vs. bottom quartile, among premenopausal women fixed effects OR (95% CI)=0.89 (0.72-1.11), among postmenopausal women with proximate collection random effects OR (95% CI)=1.29 (0.71-2.38)).

**Table 3a:**
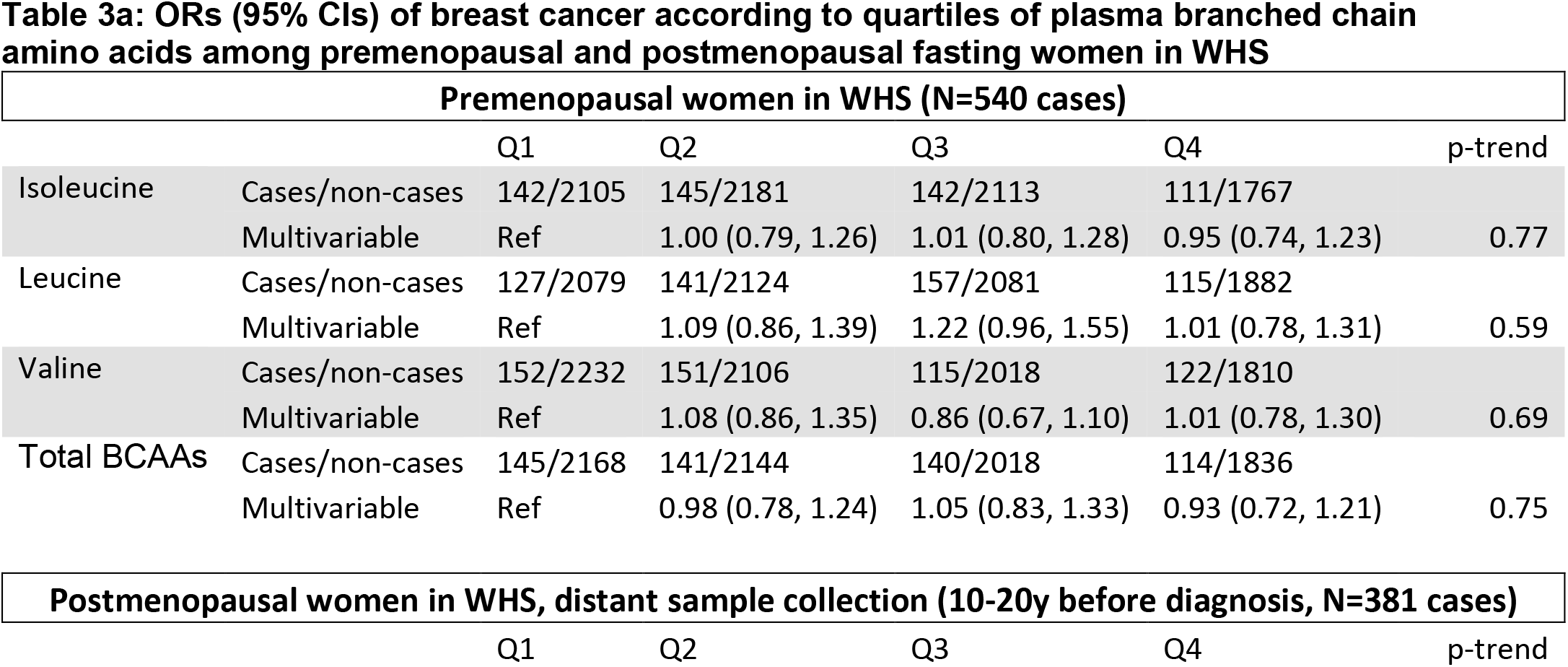
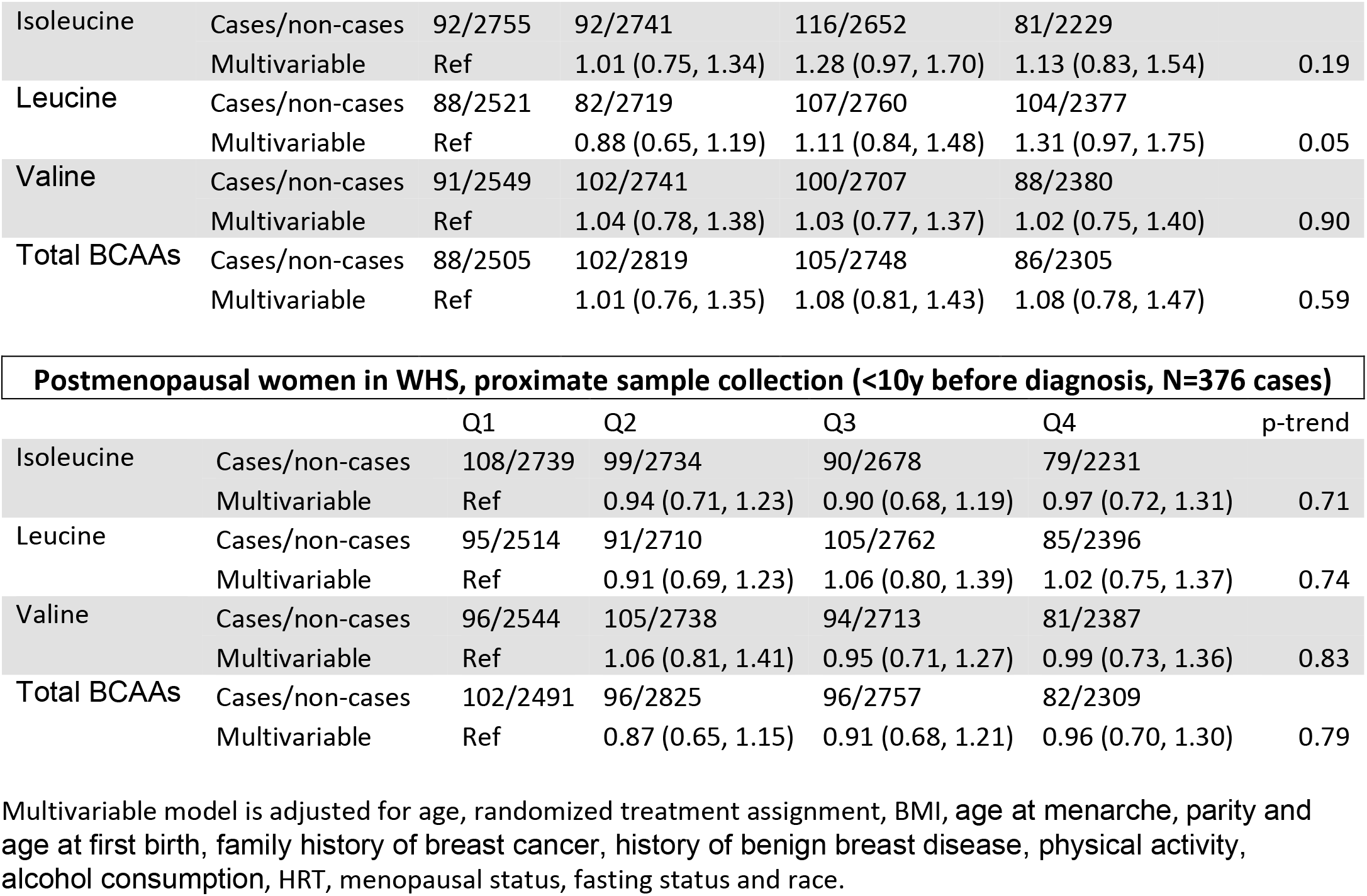
ORs (95% CIs) of breast cancer according to quartiles of plasma branched chain amino acids among premenopausal and postmenopausal fasting women in WHS

| Premenopausal women in WHS (N=540 cases) |  |  |  |  |  |  |
| --- | --- | --- | --- | --- | --- | --- |
|  |  | Q1 | Q2 | Q3 | Q4 | p-trend |
| Isoleucine | Cases/non-cases | 142/2105 | 145/2181 | 142/2113 | 111/1767 |  |
|  | Multivariable | Ref | 1.00 (0.79, 1.26) | 1.01 (0.80, 1.28) | 0.95 (0.74, 1.23) | 0.77 |
| Leucine | Cases/non-cases | 127/2079 | 141/2124 | 157/2081 | 115/1882 |  |
|  | Multivariable | Ref | 1.09 (0.86, 1.39) | 1.22 (0.96, 1.55) | 1.01 (0.78, 1.31) | 0.59 |
| Valine | Cases/non-cases | 152/2232 | 151/2106 | 115/2018 | 122/1810 |  |
|  | Multivariable | Ref | 1.08 (0.86, 1.35) | 0.86 (0.67, 1.10) | 1.01 (0.78, 1.30) | 0.69 |
| Total BCAAs | Cases/non-cases | 145/2168 | 141/2144 | 140/2018 | 114/1836 |  |
|  | Multivariable | Ref | 0.98 (0.78, 1.24) | 1.05 (0.83, 1.33) | 0.93 (0.72, 1.21) | 0.75 |

**Postmenopausal women in WHS, distant sample collection (10-20y before diagnosis, N=381 cases)**
|  | Q1 | Q2 | Q3 | Q4 | p-trend |
| --- | --- | --- | --- | --- | --- |
| Isoleucine | Cases/non-cases | 92/2755 | 92/2741 | 116/2652 | 81/2229 |  |
|  | Multivariable | Ref | 1.01 (0.75, 1.34) | 1.28 (0.97, 1.70) | 1.13 (0.83, 1.54) | 0.19 |
| Leucine | Cases/non-cases | 88/2521 | 82/2719 | 107/2760 | 104/2377 |  |
|  | Multivariable | Ref | 0.88 (0.65, 1.19) | 1.11 (0.84, 1.48) | 1.31 (0.97, 1.75) | 0.05 |
| Valine | Cases/non-cases | 91/2549 | 102/2741 | 100/2707 | 88/2380 |  |
|  | Multivariable | Ref | 1.04 (0.78, 1.38) | 1.03 (0.77, 1.37) | 1.02 (0.75, 1.40) | 0.90 |
| Total BCAAs | Cases/non-cases | 88/2505 | 102/2819 | 105/2748 | 86/2305 |  |
|  | Multivariable | Ref | 1.01 (0.76, 1.35) | 1.08 (0.81, 1.43) | 1.08 (0.78, 1.47) | 0.59 |

| Postmenopausal women in WHS, proximate sample collection (<10y before diagnosis, N=376 cases) |  |  |  |  |  |  |
| --- | --- | --- | --- | --- | --- | --- |
|  |  | Q1 | Q2 | Q3 | Q4 | p-trend |
| Isoleucine | Cases/non-cases | 108/2739 | 99/2734 | 90/2678 | 79/2231 |  |
|  | Multivariable | Ref | 0.94 (0.71, 1.23) | 0.90 (0.68, 1.19) | 0.97 (0.72, 1.31) | 0.71 |
| Leucine | Cases/non-cases | 95/2514 | 91/2710 | 105/2762 | 85/2396 |  |
|  | Multivariable | Ref | 0.91 (0.69, 1.23) | 1.06 (0.80, 1.39) | 1.02 (0.75, 1.37) | 0.74 |
| Valine | Cases/non-cases | 96/2544 | 105/2738 | 94/2713 | 81/2387 |  |
|  | Multivariable | Ref | 1.06 (0.81, 1.41) | 0.95 (0.71, 1.27) | 0.99 (0.73, 1.36) | 0.83 |
| Total BCAAs | Cases/non-cases | 102/2491 | 96/2825 | 96/2757 | 82/2309 |  |
|  | Multivariable | Ref | 0.87 (0.65, 1.15) | 0.91 (0.68, 1.21) | 0.96 (0.70, 1.30) | 0.79 |

Multivariable model is adjusted for age, randomized treatment assignment, BMI, age at menarche, parity and age at first birth, family history of breast cancer, history of benign breast disease, physical activity, alcohol consumption, HRT, menopausal status, fasting status and race.

Results among NHS/NHSII women did not change in sensitivity analyses (data not shown) among pre- and postmenopausal women separately in which we adjusted, in individual models, for BMI at the time of the blood collection instead of BMI at age 18 and weight change between age 18 and blood collection, plasma C-peptide levels, and plasma estradiol levels.

No associations were observed for individual and total BCAAs when we cross-classified BCAA levels 10 years apart. However, we observed a 3-fold increase in risk of breast cancer (multivariable OR(95%CI)=3.00(1.45-6.20)) for NHS participants with low isoleucine levels in the first blood collection but high isoleucine levels in the second blood collection (low/high) compared to participants who had low isoleucine levels in both blood collections (low/low; Table 4).

**Table 4:**
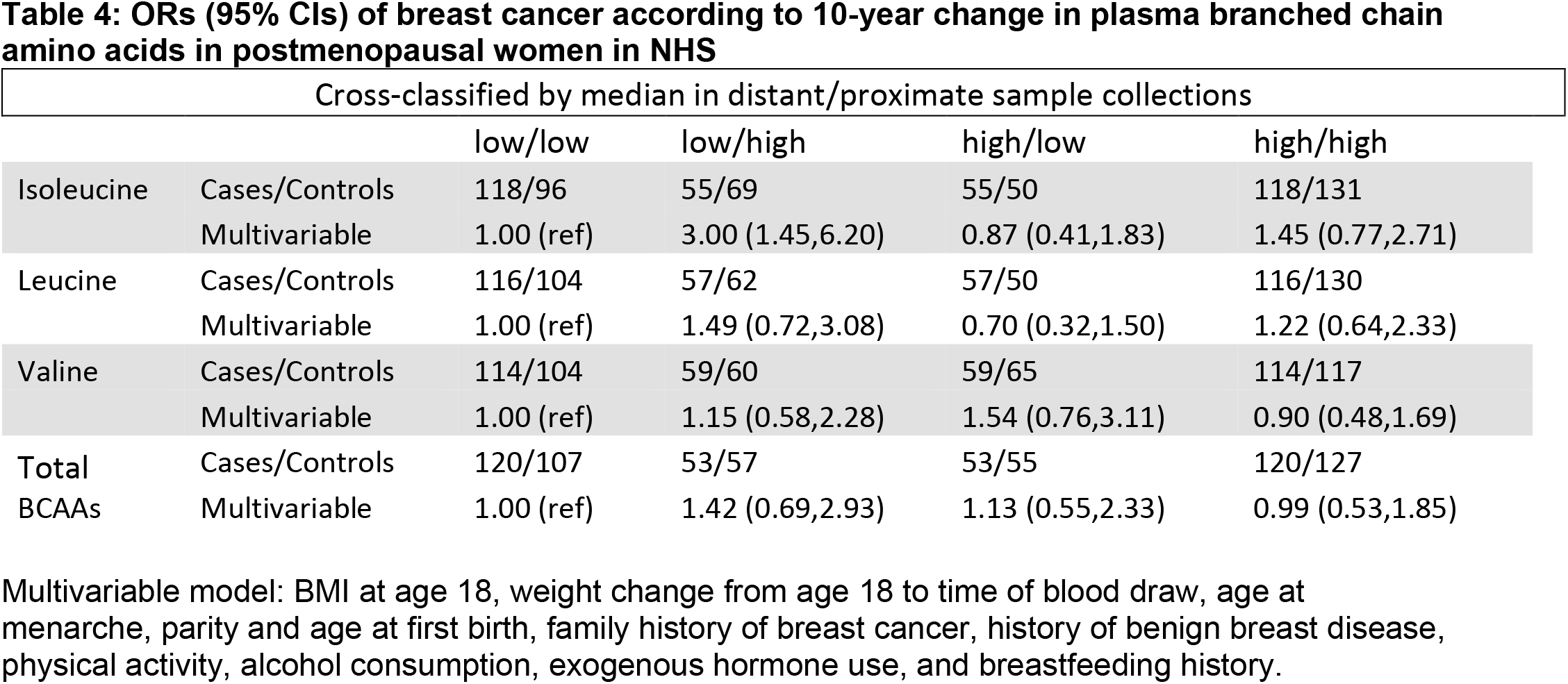
ORs (95% CIs) of breast cancer according to 10-year change in plasma branched chain amino acids in postmenopausal women in NHS.

Interactions with BMI were not significant. However, among fasting overweight/obese (BMI ≥25) premenopausal women, individual and total BCAAs were inversely associated with risk, with significant linear trends (e.g., isoleucine OR(95%CI)=0.61(0.37-1.01), p-trend=0.05; Supplementary Table 1), while no association was observed in fasting lean premenopausal women. Further, among fasting postmenopausal women positive associations with risk were observed among lean women with proximate blood collection for isoleucine, valine and total BCAAs (OR (95%CI), lean women=2.26(1.18-4.31), p-trend=0.01).

There were no significant associations between BCAA levels and breast cancer risk by ER status (Supplementary Table 2).

In breast tumor gene expression analyses, among NHSII and NHS samples separately, similar pathway activity was observed for each of the individual BCAAs. Circulating BCAA levels were associated with upregulation of mTOR signaling, interferon response, MYC, E2F and G2M targets, and DNA repair among NHSII women (68% premenopausal at blood collection) but with upregulation of estrogen response among NHS participants (all postmenopausal women; Supplementary Table 3).

## Discussion

In this prospective analysis of BCAAs and risk of breast cancer with 1,997 case-control pairs, elevated circulating BCAA levels were associated with lower risk of breast cancer among premenopausal women at blood collection but higher risk of breast cancer among postmenopausal women at blood collection with proximate (<10y before diagnosis) assessments, independent of measures of adiposity. Associations were similar across individual and total BCAAs. Both inverse and positive associations were slightly stronger with significant linear trends when we restricted to fasting samples (a significant predictor of circulating BCAA levels), which may better reflect underlying metabolic dysregulation compared with samples collected shortly after meals, when BCAA levels may be more likely to reflect recent dietary intake than long-term metabolic state^35^. No significant associations were observed when assessing distant measures of BCAAs among postmenopausal women. We did not observe interactions with BMI or heterogeneity by ER status. Associations did not validate in WHS.

BCAAs are essential nutrients that must be acquired from food or biosynthesized by the microbiome^36^. Several studies have found a positive but weak correlation between dietary BCAA intake and circulating BCAAs (r=0.11 -0.14)^37-40^. Similarly, we observed that dietary intake was a significant but fairly weak predictor of circulating levels. Diets high in animal protein, especially red meat, are associated with increased BCAA levels compared to those with predominately plant sources of protein^40-43^. We and others have shown, including a large meta-analysis, that higher intake of red meat is associated with increased risk of pre- and postmenopausal breast cancer^44,45^. In the Primary Prevention of Cardiovascular Disease with a Mediterranean Diet trial (PREDIMED), participants randomized to follow a Mediterranean diet (high intake of fruit, vegetable, grains, beans, nuts and seeds and low to moderate intake of dairy, eggs, fish and poultry) supplemented with olive oil showed significantly reduced plasma BCAA levels after one year compared to the low-fat control dietary intervention^46^. However, BCAAs were not identified as markers of dietary patterns^47^ or dietary intake^48^, suggesting that the role of BCAAs in breast cancer etiology may reflect mechanisms beyond their dietary intake.

The role of obesity in postmenopausal breast cancer is well established^49,50^, and diabetes and insulin resistance have been associated with breast cancer risk^51^. Elevated levels of circulating BCAAs are associated with obesity and insulin resistance in cross-sectional studies^52-54^, and predictive of future incidence of Type II Diabetes (T2D) in longitudinal studies^40,55^. Mendelian randomization analyses have shown that adiposity and insulin resistance have a causal effect on serum BCAA levels^56-58^, and circulating BCAAs play a causal role in the development of T2D^37^. Together, these findings emphasize that elevated BCAA levels are indicative of a dysregulated metabolism. Further, dietary BCAAs in experimental and human studies cause impaired insulin activity through upregulation of the mTOR pathway^59,60^, which has been implicated in breast carcinogenesis^61^.

Our observed opposite associations between plasma BCAAs and breast cancer risk by menopausal status parallel the associations between BMI and breast cancer risk by menopausal status, though associations with BCAAs persisted even with adjustment for BMI at age 18 and weight change or BMI at the time of sample collection. The association was also independent of plasma estradiol levels. We also observed differential associations by menopausal status between circulating BCAAs and breast tumor gene expression, with mTOR and interferon signaling, DNA repair, and E2F, G2M and MYC targets among premenopausal women, but estrogen response among postmenopausal women. Taken together, these findings suggest that BCAAs play a role in breast carcinogenesis beyond their role in obesity and insulin resistance.

Few epidemiologic studies have investigated the association of circulating BCAA levels with risk of breast cancer and only one assessed this relationship by menopausal status. Kuhn et al. observed no significant association between individual BCAAs and risk of breast cancer^11^ in a case-cohort analysis in the EPIC-Heidelberg cohort (N=362 cases, 114 pre- and 248 postmenopausal). No significant associations were observed for any of the individual BCAAs^10^ in a recent large prospective study in the EPIC cohort (N=1,624 cases, 434 pre-, 318 peri- and 872 postmenopausal). Higher levels of valine were associated with increased risk of breast cancer^12^ among pre- and postmenopausal women within the Supplementation en Vitamines et Mineraux Antioxydants study (SU.VI.MAX; N=211 cases, 129 pre- and 82 postmenopausal). Given the mix of menopausal status in these studies, it is difficult to compare these results to our findings. Similar to our results, in a nested case-control study within the PLCO cohort that examined BMI-correlated metabolites, which included valine and allo-isoleucine, (N=621 cases, all postmenopausal), higher levels of allo-isoleucine were associated with increased risk of postmenopausal breast cancer^13^. Allo-isoleucine is a byproduct of isoleucine transamination^62^. Notably, two other metabolites involved in BCAA metabolism were positively associated with risk, 2-methylbutyrylcarnitine (indicating alternative isoleucine degradation) and 3-methylglutarylcarnitine (indicating alternative leucine degradation)^13^. Sensitivity analyses adjusting for insulin resistance-related metabolites resulted in only slight attenuation of the association of BCAAs and breast cancer risk. Similarly, in the current study we observed no changes when adjusting for C-peptide, a measure of insulin production, suggesting that the role of BCAAs in postmenopausal breast cancer etiology be independent of their function in insulin resistance. In summary, results from PLCO and NHS/NHSII suggest that isoleucine and leucine may play a role in postmenopausal breast cancer although findings from WHS were not consistent. However, to which extent individual BCAAs contribute to breast cancer and how this relationship is modulated by menopausal status is not clear. Additional prospective cohort studies are needed to confirm these relationships.

Several lines of indirect evidence indicate a role of BCAA metabolism in cancer growth, progression and metastasis^63^. Amino acids represent a significant source of energy during oncogenesis^64^ with BCAA uptake being additionally upregulated in tumors^65^. Higher tissue expression of the enzymes involved in BCAA degradation was observed for several cancers^66-69^, including breast cancer^70^. Furthermore, elevation of circulating BCAAs is an early event in human pancreatic adenocarcinoma development^71^. These findings suggest that BCAA metabolism plays a critical role in cancer progression.

Our study has several strengths and limitations. We measured plasma BCAAs among a large number of pre- and postmenopausal women before diagnosis of breast cancer in two large cohort studies. We had detailed information on breast cancer risk factors, including measures of adiposity. We had limited statistical power in the analysis of ER-tumors. There were only a limited number of participants who provided two blood samples; thus, our main findings are based on one-point-in-time blood sample. However, BCAAs showed good within-person stability over 1-2 years (intra-class correlation≥0.55)^19^ as well as good within-person stability over 10 years (intra-class correlation>0.4). Metabolomics platforms differed between NHS/NHSII and WHS; NMR approaches may be more limited in measuring BCAA levels^72^. However, others showed good correlations between the two platforms and consistent associations with diabetes across platforms^73^.

In summary, we found that elevated circulating BCAA levels were associated with higher risk of postmenopausal breast cancer in NHS when assessed within 10 years of diagnosis, independent of established risk factors, including adiposity, though this finding was not replicated among predominantly postmenopausal women in WHS. Whether circulating BCAAs levels are inversely associated with risk of breast cancer among premenopausal women warrants further investigation. Reasonably powered population and experimental studies are needed to understand the mechanisms underlying this relationship. Further understanding of how circulating BCAA levels, modulated by menopausal status, potentially drive carcinogenic processes may open new directions for the development of prevention and treatment protocols. Studies investigating the predictive capacity of circulating BCAA levels could help improve current risk prediction models and help identify women at high risk.

## Data Availability

Data will be made available upon reasonable request.

## Acknowledgements

The NHS/NHSII was funded by the National Cancer Institute (R01 CA050385, UM1 CA186107, P01 CA087969, R01 CA49449, U01 CA176726, R01 CA67262). The WHS is supported by the National Institutes of Health (grant numbers CA-047988, HL-043851, HL-080467, HL-099355, and UM1 CA182913) and Dr. Mora has received institutional research grant support from the National Heart, Lung, and Blood Institute (R01HL134811 and K24 HL136852), and the National Institute of Diabetes and Digestive and Kidney Diseases (DK112940).We would like to thank the participants and staff of the Nurses’ Health Studies for their valuable contributions as well as the following state cancer registries for their help: AL, AZ, AR, CA, CO, CT, DE, FL, GA, ID, IL, IN, IA, KY, LA, ME, MD, MA, MI, NE, NH, NJ, NY, NC, ND, OH, OK, OR, PA, RI, SC, TN, TX, VA, WA, WY. The authors assume full responsibility for analyses and interpretation of these data.

